# An Integrated Genomic and Transcriptomic Analysis Reveals Distinct Molecular Features Associated with Mild-to-moderate Atopic Dermatitis

**DOI:** 10.1101/2023.04.12.23288462

**Authors:** Tu Hu, Tanja Todberg, Lone Skov, Thomas Litman, Ilka Hoof, Joel Correa da Rosa

**Affiliations:** Explorative Biology and Bioinformatics, LEO Pharma A/S; Department of Immunology and Microbiology, University of Copenhagen; Department of Dermatology and Allergy, Copenhagen University Hospital – Herlev and Gentofte; Department of Clinical Medicine, University of Copenhagen; Laboratory of Inflammatory Skin Diseases, Icahn School of Medicine at Mount Sinai, New York

**Keywords:** Atopic dermatitis, genetic risk score, transcriptome, inflammation, fibroblast, IL-34, genetic variant

## Abstract

Atopic dermatitis (AD) is a common skin disorder, characterized by impaired skin barrier function and cutaneous inflammation. The pathophysiology of AD is incompletely understood, and has considerable genetic contributions. To obtain a detailed molecular understanding of AD, we integrated the genomic, skin transcriptomic, and clinical measurements from 30 AD and 30 healthy control (HC) subjects. We found that the AD group had mild-to-moderate disease severity and only showed slightly increased genetic risk compared with HC. When comparing within the AD group, we found that the lesional skin of patients with increased genetic risk was characterized by a possible “self-protection” mechanism, including elevation of the anti-inflammatory cytokine IL-34, activation of fibroblasts, wound healing, and the complement system. We hypothesize that this mechanism may contribute to halting further progression of AD.

## Introduction

Atopic dermatitis (AD) is a common inflammatory skin disorder, characterized by skin barrier abnormalities and T-cell driven inflammation. The pathophysiology of AD is complex and incompletely understood (Weidinger and Novak, 2015). AD has a considerable genetic contribution with a heritability of 60-90% as reported in twin studies (Thomsen et al., 2007; Bataille et al., 2012; Elmose and Thomsen, 2015).

The EArly Genetic and Lifecourse Epidemiology (EAGLE) study, the most recent and by far the largest genome-wide association study (GWAS) of AD, has identified 29 loci associated with AD in the European population (Paternoster et al., 2015). The majority of the loci associated with AD are located in the noncoding region of the genome, which indicates that the genetic variants have a regulatory role, rather than directly affecting protein function. The reported SNPs all had small effect sizes (median odds ratio (OR) = 1.08, range: 1.04 - 1.61)

With the advancement of RNA-seq, transcriptomic studies have identified numerous differentially expressed genes associated with AD, and provided novel insights into AD pathogenesis (Tsoi et al., 2019; Möbus et al., 2020).

In the GENtofte Atopic Dermatitis (GENAD) cohort, we performed genomic, skin transcriptomic, and extensive clinical measurement on all AD and healthy control (HC) subjects, hereby generating an AD disease signature, consisting of many genes associated with activated inflammatory response and skin barrier perturbations (Hu et al., 2022). In the current study, we intend to integrate the genomic and the skin transcriptomic measurement from the same patient group to identify unique skin transcriptome features associated with AD genetic risk, and potentially, reveal the genome regulatory mechanisms in AD.

## Materials and methods

### Study cohort

The GENtofte Atopic Dermatitis (GENAD) cohort included 30 AD and 30 healthy control (HC) subjects with Northern European ancestry. The subjects were recruited between Apr 2018 and Nov 2019 at the outpatient clinic of Gentofte Hospital, Denmark. AD patients were diagnosed according to the Hanifin and Rajka criteria (Hanifin and Rajka, 1980). The patients were restricted from systemic anti-inflammatory treatment > 4 weeks and from local treatment > 2 weeks before every visit. Patients with contact allergies, malignancies, infections, and receiving immunomodulatory therapies were excluded. HCs fulfilled the same eligibility criteria as the patients with AD, except for the AD-specific criteria.

### Clinical measurement

AD patients were assessed by the physicians for disease severity, including eczema area and severity index (EASI), Scoring atopic dermatitis (SCORAD), objective SCORAD (oSCORAD), and age of onset. Total IgE was measured from the blood. HC subjects were matched in gender, age, education, BMI, and skin Fitzpatrick scale.

### Genomic data

Genomic information was profiled by Illumina Infinium Omni5Exome-4 BeadChip array (Illumina Inc., San Diego, CA) from the blood. The quality of the SNP data was checked using the imputation preparation and checking tool from the McCarthy Group (v4.2.7, https://www.well.ox.ac.uk/~wrayner/tools/) for strand, ID names, positions, alleles, ref/alt assignment. Further, the SNPs were imputed using the Michigan Imputation Server (v1.6.5). The pre-phasing step for the imputation process was performed with Eagle v2.4. Minimac4 was used for imputation with the HRC r1.1 2016 European population as reference samples. Only imputed data that fulfilled the following criteria were included in analyses: Minor allele frequency ≥ 1%, p value for Hardy-Weinberg equilibrium ≥ 1e-05, genotype and individual call rates ≥ 90%. Data cleaning, processing, and association analyses were performed in plink v1.90 (Chang et al., 2015).

### Atopic dermatitis genetic risk score

We calculated the genetic risk score (GRS) of the individuals in the GENAD cohort based on the reference GWAS data Paternoster et al., 2015, with the summary statistics retrieved from GWAS Catalog (study accession GCST003184). We mapped the imputed SNPs from our study and the reference study, which resulted in 25 matched SNPs. AD GRS was calculated based on the 25 matched SNPs, by summing the product of the effect size (β_*i*_) and dosage (*N*_*i*_) according to Igo et al., 2019, with details shown as follows: 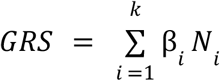, where β_*i*_: the effect size of SNP *i* reported in the reference GWAS, *N*_*i*_ : the dosage (0, 1, 2) of SNP carried by an individual in the GENAD cohort. GRS ranges between 0 - 4.2, by assuming an individual carries none to all the effect SNPs. The information of the SNPs is shown in **Table S1 (SNP info)**.

### Skin transcriptome data

The transcriptomic profiling was performed by deep RNA-seq from the lesional (LS) and matched non-lesional (NL) skin from AD patients, and site-matched healthy skin from HC. We only included baseline measurement in the analysis. In total, the expression of 14759 genes (including both protein coding and non-coding genes, expressed with a mean of more than 3 counts per million (cpm) across all samples) from 148 samples were used for the analysis. Details of sample collection and data curation were reported previously (Hu et al., 2022).

### Statistics and data analysis

We tested the differences in clinical measurements of AD (EASI, SCORAD, oSCORAD, blood IgE) between three AD genetic risk groups (low, medium, high) by ANOVA, the differences between the high and low AD genetic risk group (high vs. low) by student’s t-test, and the trend of clinical measurements when AD genetic risk increases by Jonckheere-Terpstra test. For early onset (≤2 years), the test was performed by Fisher’s exact test. We also calculated the correlation coefficients between the AD clinical measurements, and GRS by Spearman correlation.

We performed statistical and data analysis in R 4.1.2 (R core team, 2013). The trend of clinical measurement change according to GRS was assessed by Jonckheere-Terpstra test function in DescTool (Signorel, 2022). Differential gene expression analysis was performed by DESeq2 (Love et al., 2014). The pathway enrichment analysis was performed by fgsea (Sergushichev, 2016) including all genes. The rank of each gene was determined by -log10p · log2 fold change. Cell deconvolution was performed by R package MuSiC (Wang et al., 2019) and using GSE147424 (He et al., 2020b) as the reference. We have documented our data analysis pipeline in the GitHub repository (https://github.com/tuhulab/multiomics-ad-genome-a4086).

To interrogate the interaction between AD genetic risk and gene expression, we employed two-parameter logistic Bayesian models to predict the probability of developing AD (*P*), by AD genetic risk (*Z*), the gene expression (*X*), and the interaction between the two. In this analysis, we only included the genes that were identified to be differentially expressed when comparing the LS skin from patients with high *vs*. low AD genetic risk.

We formulate a Bayesian two-parameter logistic regression model as follows:

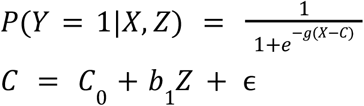

> where, *X*: log2 transformed transcript cpm
>
> *Z*: dosage {0, 1, 2} of the effect allele, or genetic risk score (0 - 4.2)
>
> *Y*: phenotype (1 for AD; 0 for HC)
>
> *C*: discriminative threshold in gene expression
>
> *C* : discriminative threshold in gene expression according to dosage 0
>
> *b* : increase in discriminative threshold caused by 1 unit increase in dosage
>
> *g*: smoothness of logistic curve (The smoother, the worse)
>
> ∈: Gaussian noise with mean 0 and variance 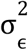

We estimated parameters *g, C*_0_, *b* and 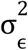 using Markov Chain Monte Carlo simulation by rjags (Plummer et al., 2022). The priors (*C*_0_ = *median*(*X*), *g* = 1, *b*_1_ = 1) were determined by checking the posterior distribution of simulated data. We deemed a model as predictive, if the highest density interval (HDI) of the logistic curve smoothness *g* posterior distribution was either in the positive (+: gene is over-expressed in AD) or in the negative range (-: gene is under-expressed in AD).

We considered that the interaction between the genetic risk and gene expression was significant, if the HDI of the threshold change (*b*_1_) posterior distribution was either in the positive (+: threshold increased) or in the negative range (-: threshold decreased).

## Results

### AD patients were characterized by mild-to-moderate disease severity

The AD patients from the GENAD cohort were characterized by mild-to-moderate AD severity, with EASI (4.2 ± 5.7; 0.2 - 22.3), SCORAD (33.5 ± 16.5; 10.8 - 67.6), oSCORAD (25.5 ± 13.5; 7.8 - 59.6) at the baseline. The above values were reported as “mean ± standard deviation; minimal - maximum value”, with individual data points reported in **Table S1**. Most of the AD patients (24 out of 30) were characterized by the intrinsic phenotype (total blood IgE ≤ 150). 19 patients had early onset (≤ 2 years), 11 had late onset AD.

### The genetic risk score of AD was low-to-medium, and associated with the clinical measurement of disease severity

After quality filtering, 933,751 SNPs were left for SNP imputation, resulting in 7,590,170 SNPs for further analysis.

We first performed an association study between the SNPs and AD, although we did not expect any significant (p < 5 × 10^−8^) SNPs to be detected due to the small sample size (n=30 for each group). The results showed that none of the SNPs had a genome-wide significant association with AD, as illustrated in the Manhattan plot (**Figure S1**).

We examined the SNPs with the highest odds ratio (OR), and observed rs6696218 to have the highest OR (21.45) with p = 8.61e-05, the effect size of which was overestimated. This genetic mutation is located in LYST, previously showing an association with markedly elevated IgE and severe atopic dermatitis in a case report for a 6-year-old male patient (Chin et al., 2021). Furthermore, we checked the SNPs showing genome-wide significant association with AD as reported in Paternoster et al., 2015, and identified the most significant SNPs to be rs61175833 (p=1.947e-05), which is located in the intergenic region with the nearest protein coding gene being SLC1A3.

Next, we calculated GRS for all individuals from the GENAD cohort study. After matching our imputed SNPs with the SNPs showing genome-wide significant association with AD in the reference GWAS study by Paternoster et al., 25 SNPs were left for further analysis. We calculated the GRS, using the effect size (β) reported in Paternoster et al., 2015. The individual data points are reported in **Table S1**.

We compared the GRS between the AD and HC group. As shown in **Figure 1**, the AD group showed a slightly increased mean and larger variation of GRS compared with the HC group (1.77 ± 0.41 *vs*. 1.66 ± 0.25, *p* = 0.23). To increase the resolution of the analysis, we decided to stratify AD patients into three AD genetic risk groups according to their GRS (low: <1.5, medium 1.5 - 1.98, high > 1.98).

**Figure 1.**
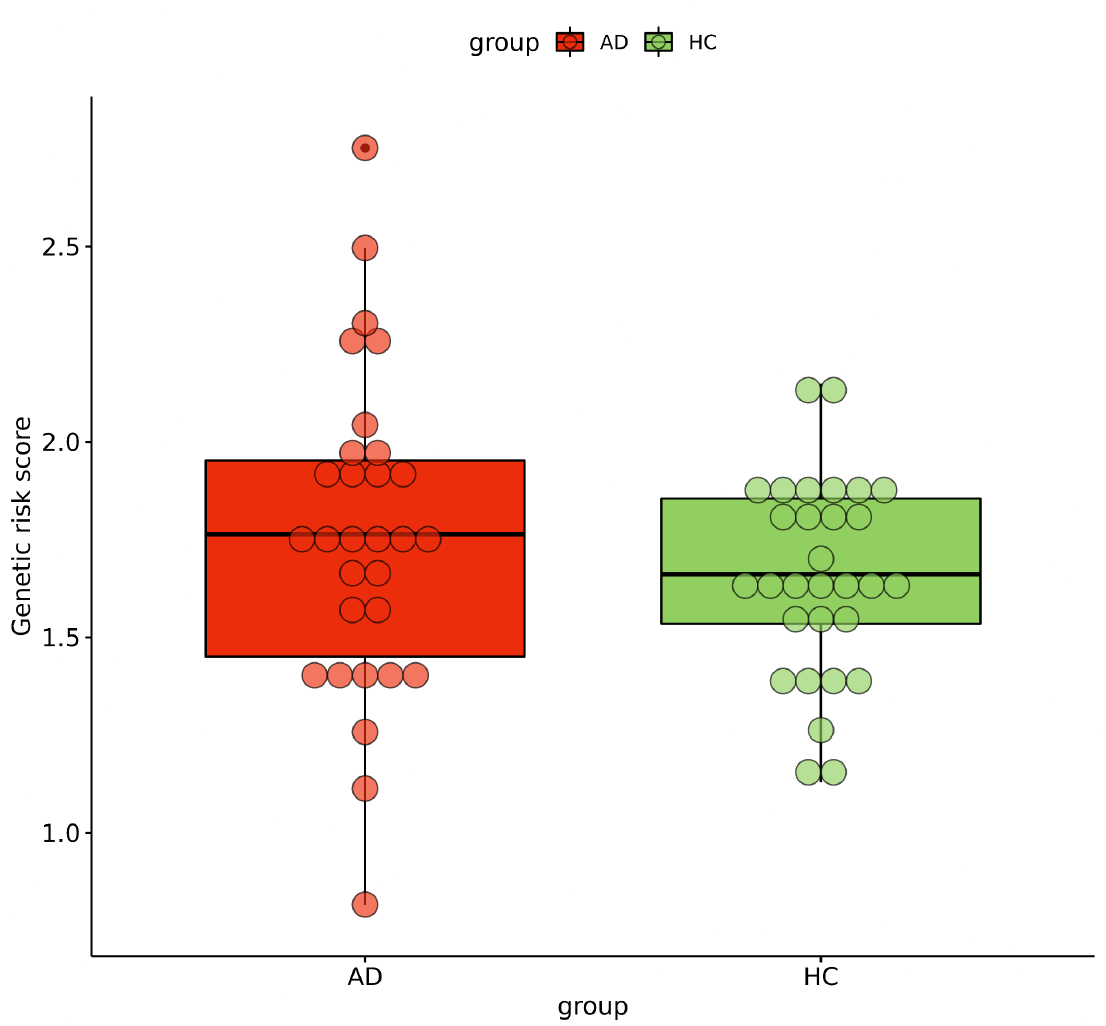
Boxplot showing the genetic risk score of AD and HC group in the GENAD cohort. The genetic risk score was calculated based on 25 SNPs (**Table S1**). The AD group showed a higher mean and variance compared with the HC group. However, the difference in GRS between the two groups was not statistically significant (*p* = 0.23, Student’s t-test).

To compare the clinical characteristics of AD patients with different genetic risk, we performed statistical testing for SCORAD, oSCORAD, EASI, IgE, and age of onset in three different AD genetic risk groups (low, medium, and high). We observed that patients in the higher AD genetic risk group were characterized by more severe AD clinical measurements and a higher probability of earlier onset. As shown in **Table 1**, the average AD disease severity scores (SCORAD, oSCORAD, EASI), total blood IgE, and the proportion of early onset all showed increasing trends, when AD genetic risk increases from low, medium, to high, although most of the statistical tests between the clinical measurements and genetic risk did not show significance. The only significant trends were observed for EASI and the genetic risk (*p* = 0. 01). The correlations between the disease severity scores and GRS are visualized in **Figure S2**. In summary, we found GRS to be able to reflect the clinical severity measurements of AD patients that were characterized by mild-to-moderate disease.

**Table 1.**
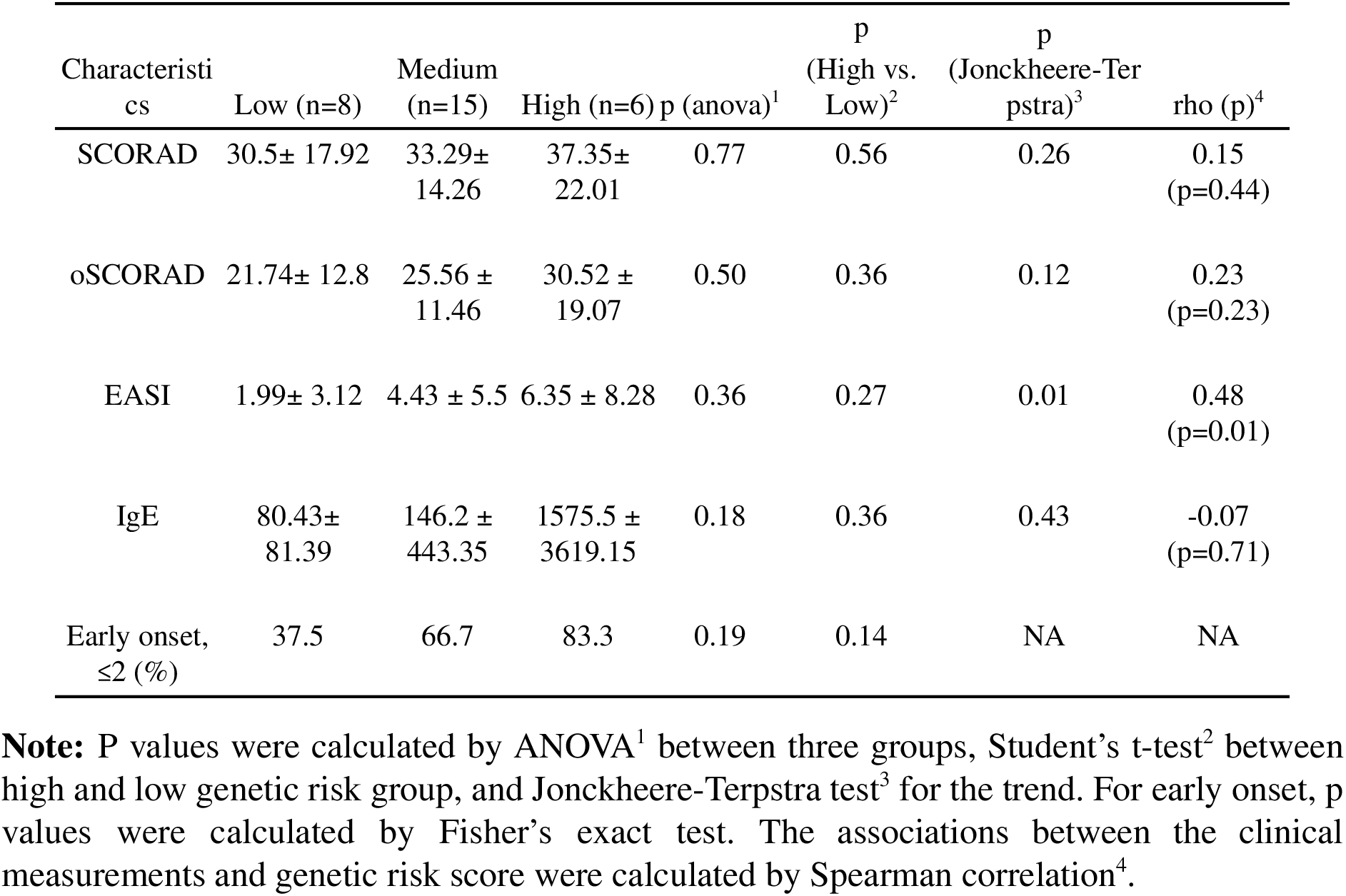
Clinical measurements of AD and correlation with AD genetic risk

### Patients with higher AD genetic risk were characterized by activated fibroblast and extracellular matrix activities in lesional skin

We studied the skin transcriptome of patients from different AD genetic risk groups. As shown in the PCA plot (**Figure 2a**), the skin transcriptome was mainly separated by the tissue state (LS/NL/HC). Most of the LS skin can be separated from HC and NL skin, but HC and NL cannot be separated. Thus, we performed separate PCA analyses for the LS (**Figure 2b**) and NL group (**Figure 2c**), but we were not able to separate skin samples based on the genetic risk of the patients in the PCA plots.

**Figure 2.**
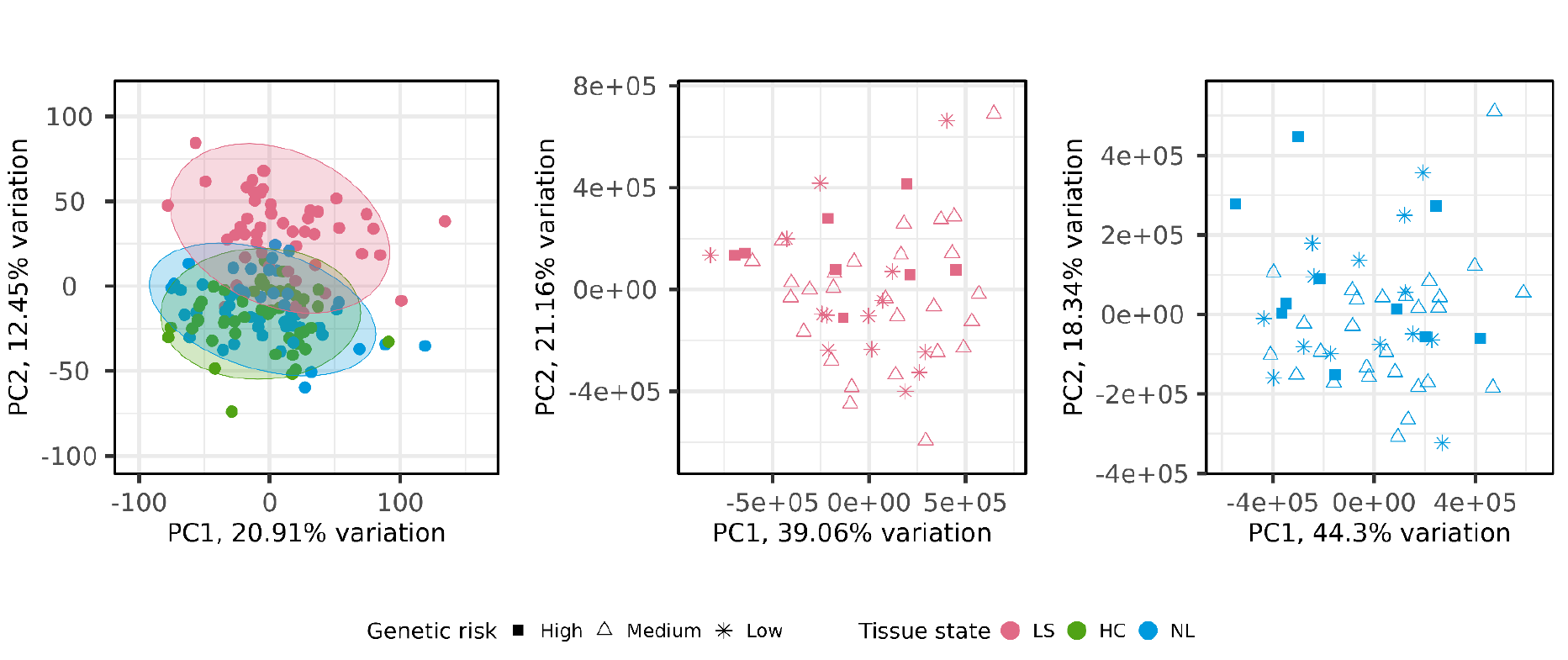
PCA plot of skin transcriptome. Genes with top 50% variance (*n* = 7350) across samples were included. (a) samples representing all three tissue states were included (LS: lesional, NL: non-lesional, HC: healthy control); (b) only LS skin; (c) Only NL skin. The shape of the symbols represents the genetic risk of the patients.

We performed differential gene expression analysis to identify the genes that show an association between expression level and AD genetic risk, with results shown in **Table S2**. We detected 58 (high *vs*. low), 46 (medium *vs*. low), and 23 (high *vs*. medium) genes to be differentially expressed (absolute log2 fold change > 1, adjusted p value < 0.05) in LS skin; 14 (high *vs*. low), 2 (medium *vs*. low), and 6 (high *vs*. medium) genes to be differentially expressed in NL skin. We only observed limited overlap between the differentially expressed genes (DEGs) between contrasts. The direction of differential expression was the same.

The comparison between LS skin in the high *vs*. low AD genetic risk group generated the largest number of differentially expressed genes. As shown in **Figure 3**, the samples from low and high risk patients can be separated based on unsupervised hierarchical clustering of 58 differentially expressed genes. Many of these genes have previously been reported to be associated with AD, with functions in maintaining skin physiological functions (wound healing and skin barrier), protein and lipid metabolism, and immune response.

**Figure 3.**
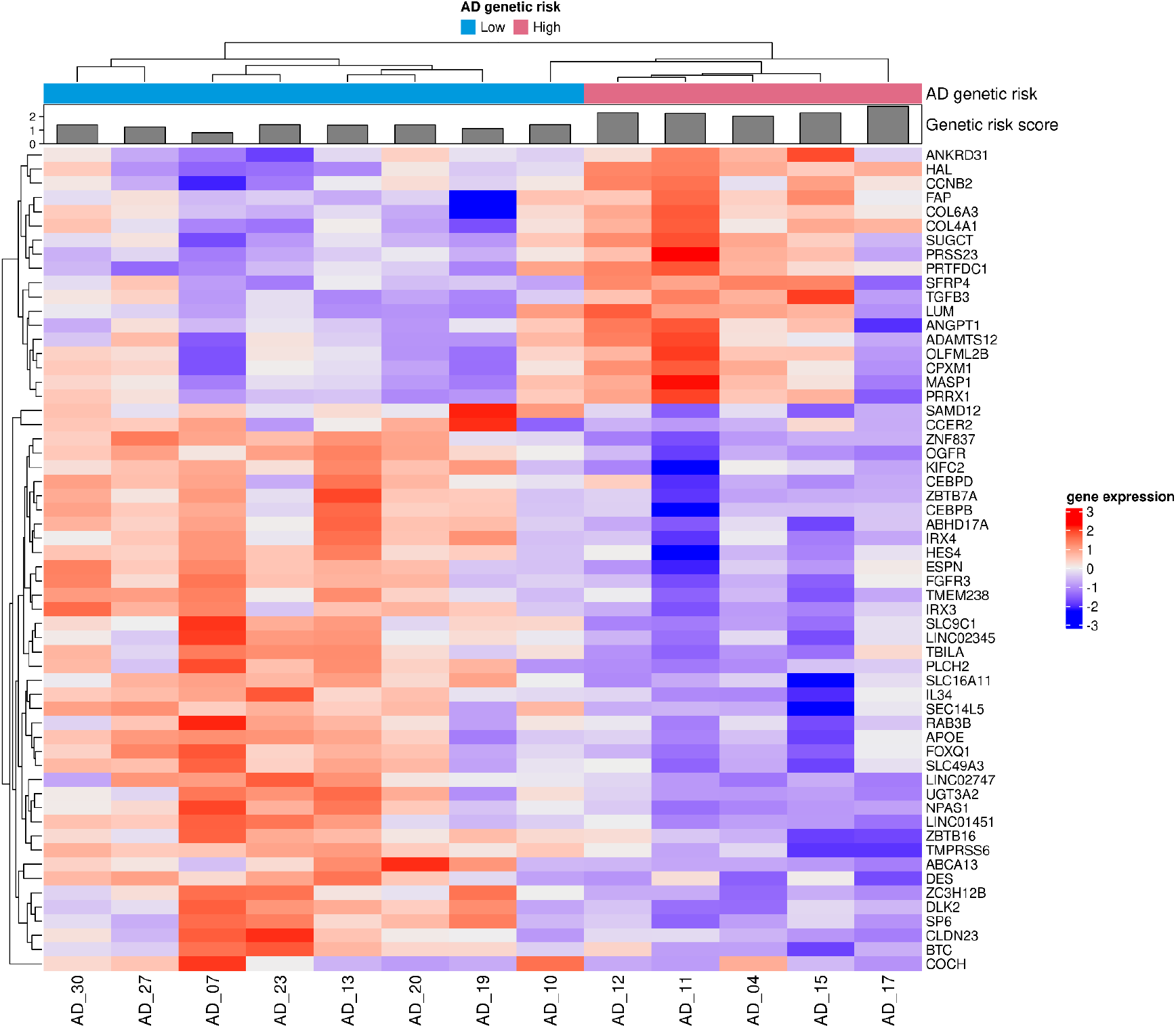
Heatmap based on 58 differentially expressed genes (high vs. low genetic risk of atopic dermatitis) in lesional skin. Each column represents a sample, named by subject ID. Each row represents a gene. AD genetic risk group (Low/High), and genetic risk score (gray bars) are shown at the top. The gene expression is z-scaled.

In the LS skin of patients with AD high genetic risk, we detected upregulation of collagen coding genes (COL4A1/6A3), genes regulating fibroblast growth (TGFB3, FAP, PRSS23), wound healing (PRRX1, ANGPT1), Wnt signaling (SFRP), extracellular matrix (LUM), histidine metabolism (HAL), and protein metabolism (CPXM1). We also detected genes downregulated in LS skin in patients with high AD genetic risk compared with low genetic risk, including lipid metabolism regulators (SLC9C1/16A11/49A3), NPAS1 (neural and nerve system), zinc-finger genes (ZNF837), skin development and scarring (SP6, OGFR), extracellular matrix (COCH, BTC, CCER2, CLDN23, TMPRSS6), melanoma suppressor gene (FOXQ1), and several lincRNAs. Interestingly, we found downregulation of UGT3A in LS skin from the high AD risk patient group. We previously reported UGT3A to have the highest fraction of variance explained by AD among all genes for the same population cohort (Hu et al., 2022). UGT3A has been proposed to maintain skin homeostasis, but its biochemical mechanism in skin physiology has not yet been elucidated. We also detected genes associated with immune response, including upregulation of MASP1 (complement activation), and downregulation of IL34. Notably, ZBTB7A and FOXQ1 are cancer suppressors. Their downregulation in LS skin from AD patients with high genetic risk might indicate that these patients have a higher risk to develop cancer compared with low AD risk patients.

We further included LS samples of patients from all genetic risk groups (low, medium, and high). As illustrated in **Figure 4**, a clear trend of differential expression of these genes was observed.

**Figure 4.**
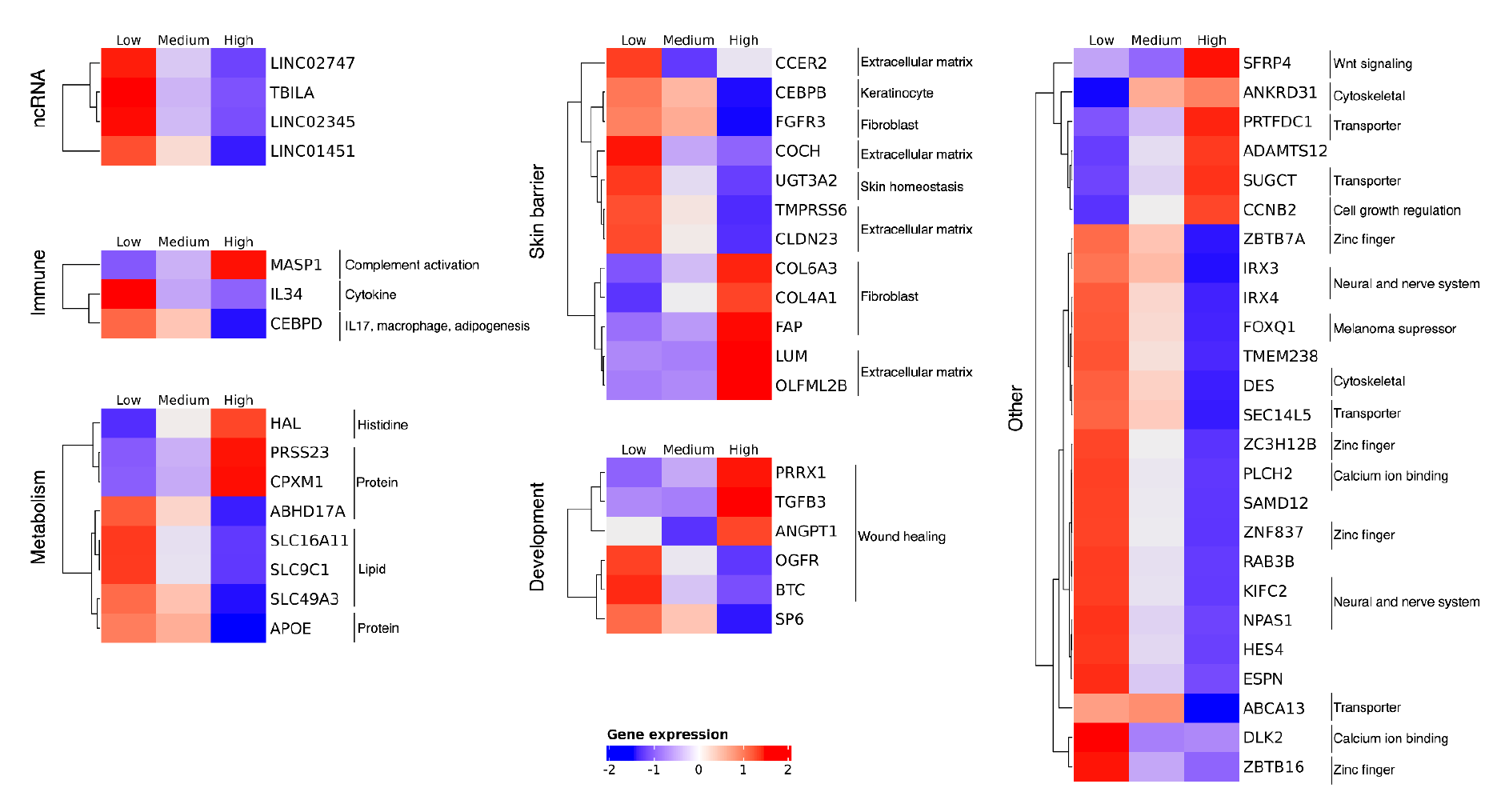
Heatmap showing the trend of differentially expressed genes in three atopic dermatitis genetic risk groups (low, medium, high). The genes are grouped by their function in skin.

We performed functional enrichment analysis of genes the expression of which was associated with AD genetic risk (high *vs*. low, high *vs*. medium, and medium *vs*. low). The results revealed that LS skin from patients with high genetic risk showed enriched signal in pathways related with extracellular matrix, collagen binding, and epidermal stasis, as shown in **Figure 5**.

**Figure 5.**
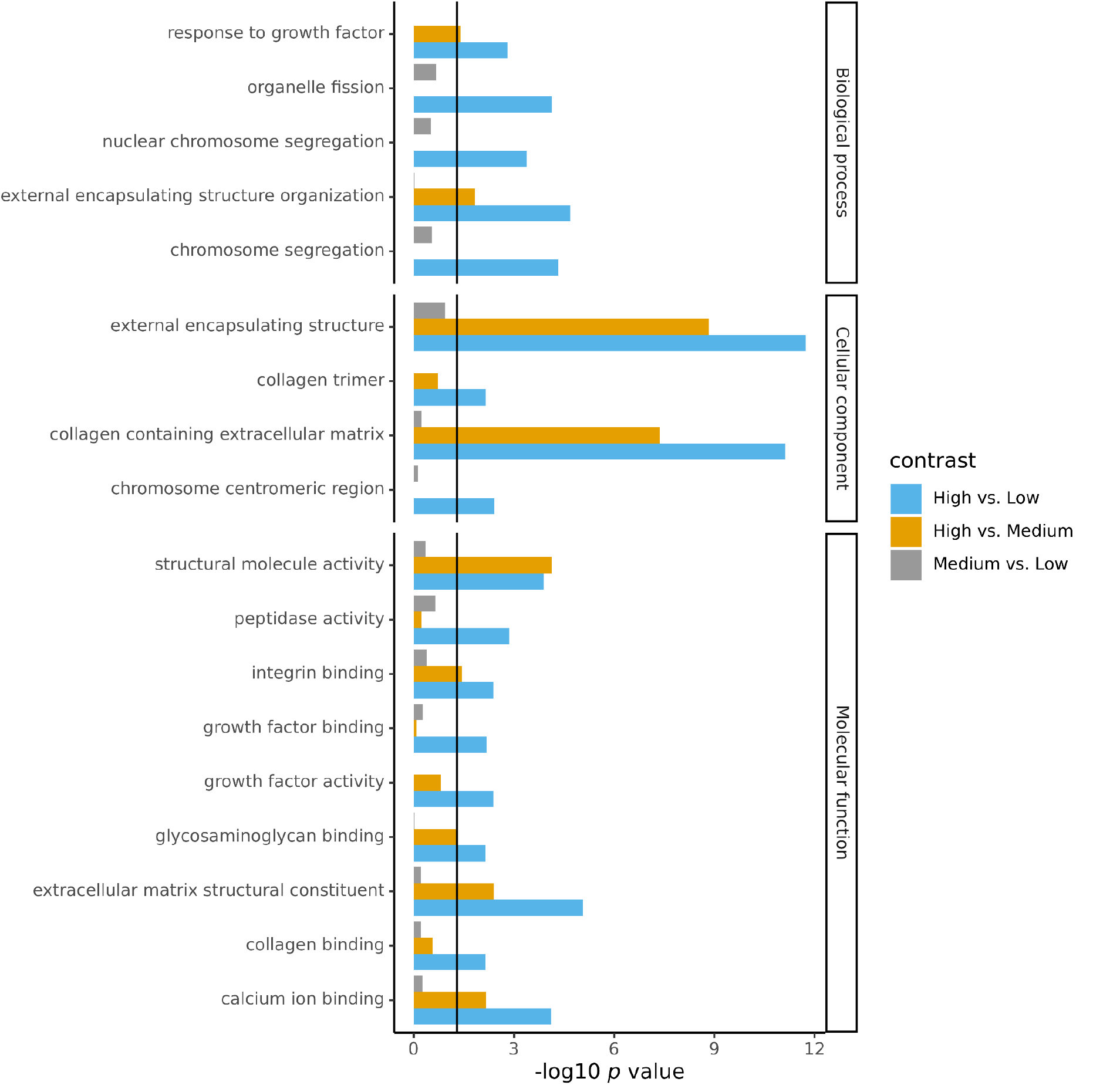
Functional enrichment analysis of atopic dermatitis lesional skin from patients with different genetic risks. The vertical line (black) shows the 0.05 significance level.

We performed cell type deconvolution analysis to identify the cell type composition change in skin from patients with different AD genetic risks. We observed that the LS skin from patients with high genetic risk showed increased proportions of fibroblasts (**Figure 6**), which confirmed our findings at the cell composition level.

**Figure 6.**
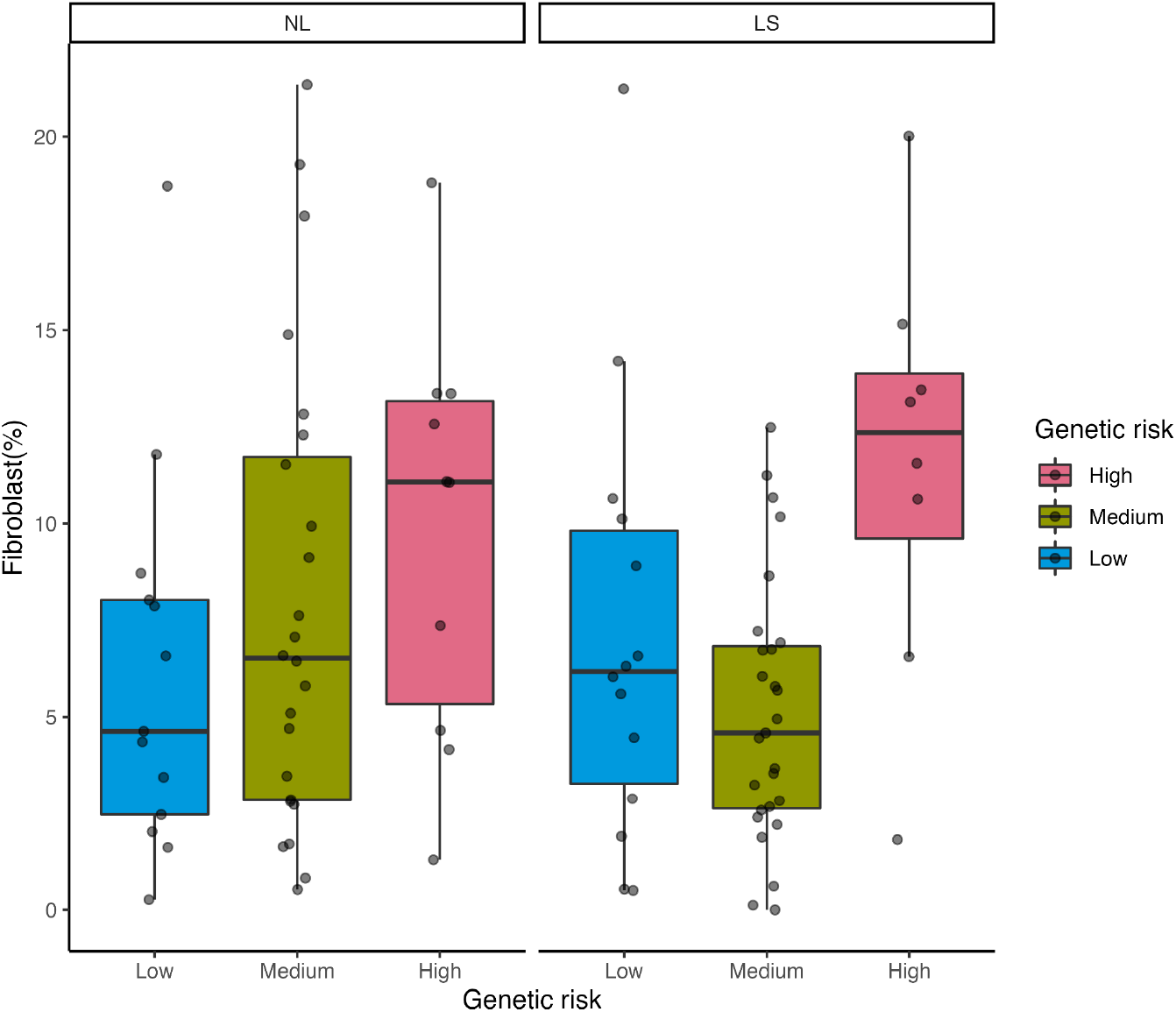
Proportion of fibroblasts in different tissue states and genetic risk groups.

### Identification of the interactions between gene expression and genetic risk of AD

To interrogate the interaction between AD genetic risk and gene expression, we employed two-parameter logistic Bayesian models to predict the probability of developing AD (*P*) by AD genetic risk (*Z*), the gene expression (*X*), and the interaction between the two.

We first used GRS as the genetic risk in the model and fitted 58 models in total, to test the combination of GRS and gene expression in predicting AD. In each model, we predicted the probability of developing AD for a subject (*P*) by the GRS (*Z*) of a subject, and the log2 transformed expression of one gene (*X*).

41 out of the 58 models showed predictive power, but it was all driven by the gene expression (*X*) alone, with none of the interactions between GRS and gene expression showing significance. Additional analyses were performed to confirm this finding (#supp-text-confirm-GRS-prediction). The limited predictive power of genetic risk is probably due to the fact that the GENAD cohort was characterized by mild-to-moderate disease severity, and low-to-moderate GRS.

Supplementary text: #supp-text-confirm-GRS-prediction

We performed additional analyses to confirm that AD genetic risk score (GRS) has limited predictive power for AD in our study.

We first checked the predictive power of the 58 genes that were differentially expressed when comparing the LS skin from patients with high *vs*. low genetic risk. Logistic regression curves were fitted to predict AD outcomes (*Y*, AD: 1; HC: 0) using log2 transformed gene expression (*X*) of all detected genes. For each logistic regression model, we predicted *Y* by *X*, then calculated the area under the curve (AUC). Then, we compared the AUC of the 58 genetic risk associated genes with the rest of the genes. We found that these 58 genes significantly outperformed the rest of the genes in predicting AD (*D* = 0. 29, *p* = 10^−4^, K-S test).

Next, we introduced AD GRS (*Z*) into the model. We found that neither additive models (*Y*∼ *Z* + *X*), nor interactive models (*Y* ∼ *Z* + *X* + *Z* * *X*) showed significant improvement in predicting AD.

Next, we evaluated the interactions between the SNPs underlying AD GRS (*n* = 25) and genes showing genetic risk specific expression pattern (*n* = 58). The interaction was modeled by similar logistic Bayesian models as described previously for GRS modelling, where, instead of the GRS, the effect allele dosage (*Z*) was included. Out of 1450 combinations between SNP dosage and gene expression, our models were able to identify 58 significant interactions between SNP and gene expression, as shown in **Figure S3**. While the rest of the interactions were non-significant, as evidenced by the observation that the highest density interval (HDI) of the *b*_1_ posterior distribution included zero. We were not able to find any significant interactions between gene expression and rs61813875 (1q21.3, the epidermal differentiation region where FLG is tagged) in our data, although it was reported as the most significantly associated SNP with AD in Paternoster et al., 2015. Ranking the genes by their b1 in the models with rs61813875, the most upregulated gene is DES (Chr 2, b1 mode = - 0.42, b1 HDI: -3.27 - 1.44), and the most downregulated gene is COL4A1 (Chr 13, b1 mode = 0.39, b1 HDI: -0.73 - 1.38).

When comparing our SNP-gene expression interaction results with the triangulation study (Sobczyk et al., 2021), we were able to identify genes from the same pathway, although the interactions were not observed for the same SNPs. We identified the following interactions: SLC16A11-rs2212434 (OR=0.37), SLC16A11-rs7927894 (OR=0.26), ZBTB7A-rs4713555 (OR=3.27), ZBTB16-rs7927894 (OR=0.26), KIFC2-rs4713555 (OR=2.84), KIFC2-rs7927894 (OR=2.84), RAB3B-rs2212434 (OR=0.12), RAB3B-rs7927894 (OR=0.13).

## Discussion

In the current study, we integrated genomic and skin transcriptomic measurements on the same study cohort and identified a distinct gene expression profile associated with higher genetic risk in mild-to-moderate AD. Unlike the canonical AD transcriptome signature, which was dominated by T_H_2 driven inflammatory response and impaired skin barrier signature, we found that LS skin from patients with increased AD genetic risk showed elevated IL34, an anti-inflammatory cytokine, activated fibroblast, extracellular matrix, wound healing, complement system, as well as a decreased expression of FOXQ1, a cancer suppressor in the Wnt signaling pathway.

We hypothesize that due to increased genetic risk, a “self-protection” program, including skin barrier repair, anti-inflammatory and antibacterial pathways, could be activated in the LS skin and thus contribute to halting the progression of AD.

Currently, the role of such a “self-protection” program has not been clarified in AD, although several studies have shed light on it. For example, a previous scRNA-seq study of AD has suggested that fibroblast subpopulations (COL6A5^+^ COL18A1^+^) may play a cross-talking role between dendritic cells and T cells to orchestrate T-cell migration and Th2/Tc2 polarization in AD (He et al., 2020a). Lefèvre-Utile et al. have reported an AD endotype, which was characterized by high negative immune regulation and skin barrier repair and associated with less severe AD and low *Staphylococcus aureus* colonization (Lefèvre-Utile et al., 2022). This study also points out the primary limitation of our study; that due to small sample size and a homogeneous study population, an AD endotype seems to be dominant in our study.

It is worthy to note that we identified differentially expressed Wnt signaling pathway members (FOXQ1, SFRP4) in AD patients with higher genetic risk. FOXQ1 has a complex and controversial role in different cancers. In colorectal cancer, FOXQ1 has been reported to be over-expressed (Kaneda et al., 2010); but in breast cancer, its lower expression is associated with poor survival (Elian et al., 2021, 1). In skin, evidence has shown FOXQ1 to be able to suppress melanoma (Bagati et al., 2017, 1). A recent study has reported that IL-4 drives the activity of FOXQ1, and FOXQ1 is over-expressed in monocytes in blood from AD patients (Ovsiy et al., 2017).

IL-4 and the IL-4 receptor are important therapeutic targets for AD and other inflammatory disorders, including inflammatory bowel disease, asthma, arthritis, and rhinitis. However, currently, the link between AD and risk of cancer is inconclusive.

Our Bayesian models were able to identify a few significant interactions between the SNP and gene expression to increase the predictive power for AD. These significant interactions appear to overlap with the triangulating study by (Sobczyk et al., 2021), but we cannot conclude the genes to be causal at the SNP loci. Data from other AD studies should be used to validate our findings.

## Data Availability

RNA sequencing data are available in Gene Expression Omnibus under the accession GSE193309. Genetic risk score is available in the supplementary table.

https://www.ncbi.nlm.nih.gov/geo/query/acc.cgi?acc=GSE193309

## Supplementary Figures

**Figure S1.**
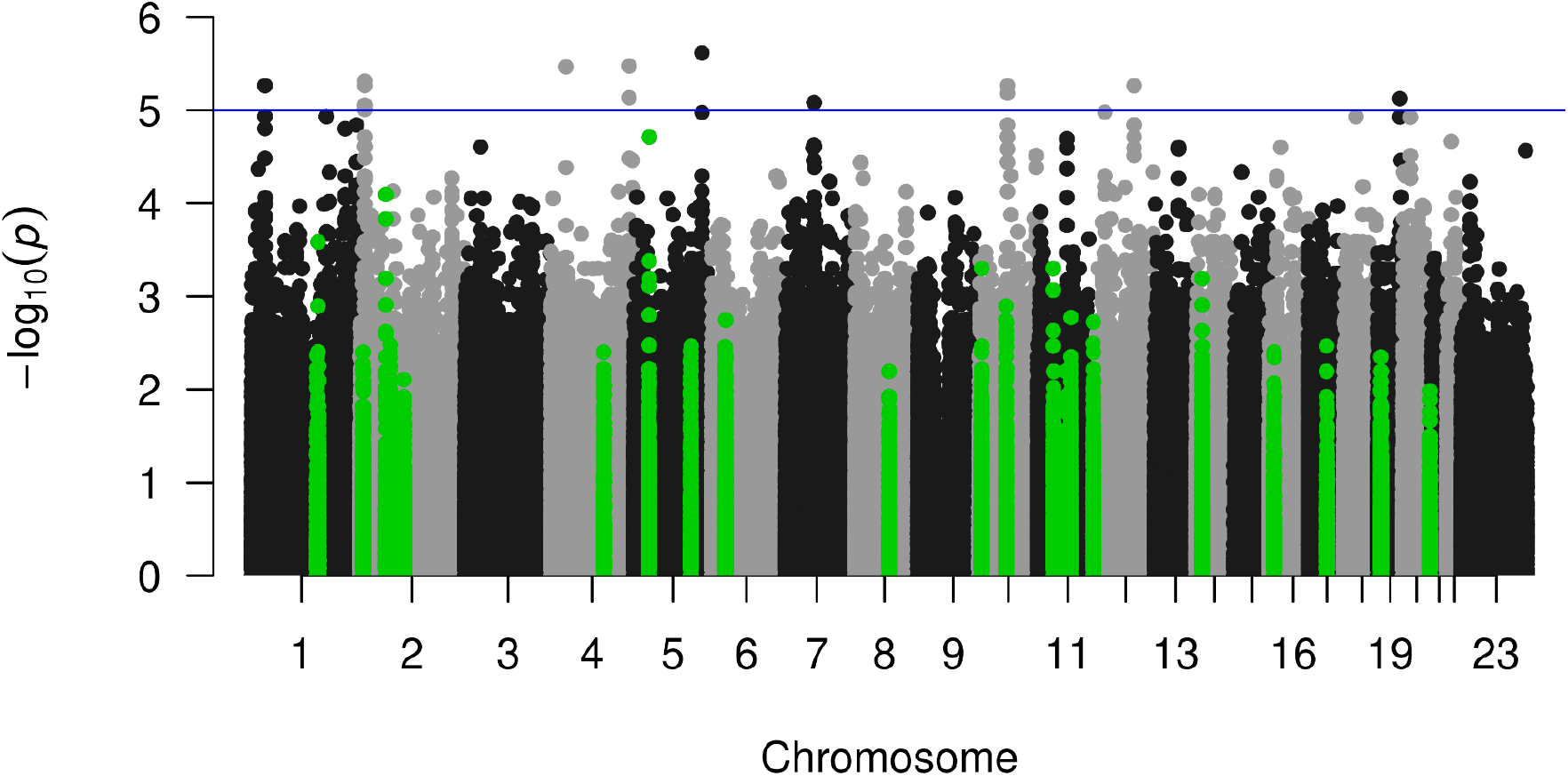
Manhattan plot showing the association between the SNP and atopic dermatitis in the GENAD cohort (30 AD vs. 30 HC). The analysis was based on imputed SNPs after quality filtering (n = 5,663,014). The green dots are ±1M bp of SNPs showing genome-wide significance in the European ancestry from (Paternoster et al., 2015).

**Figure S2.**
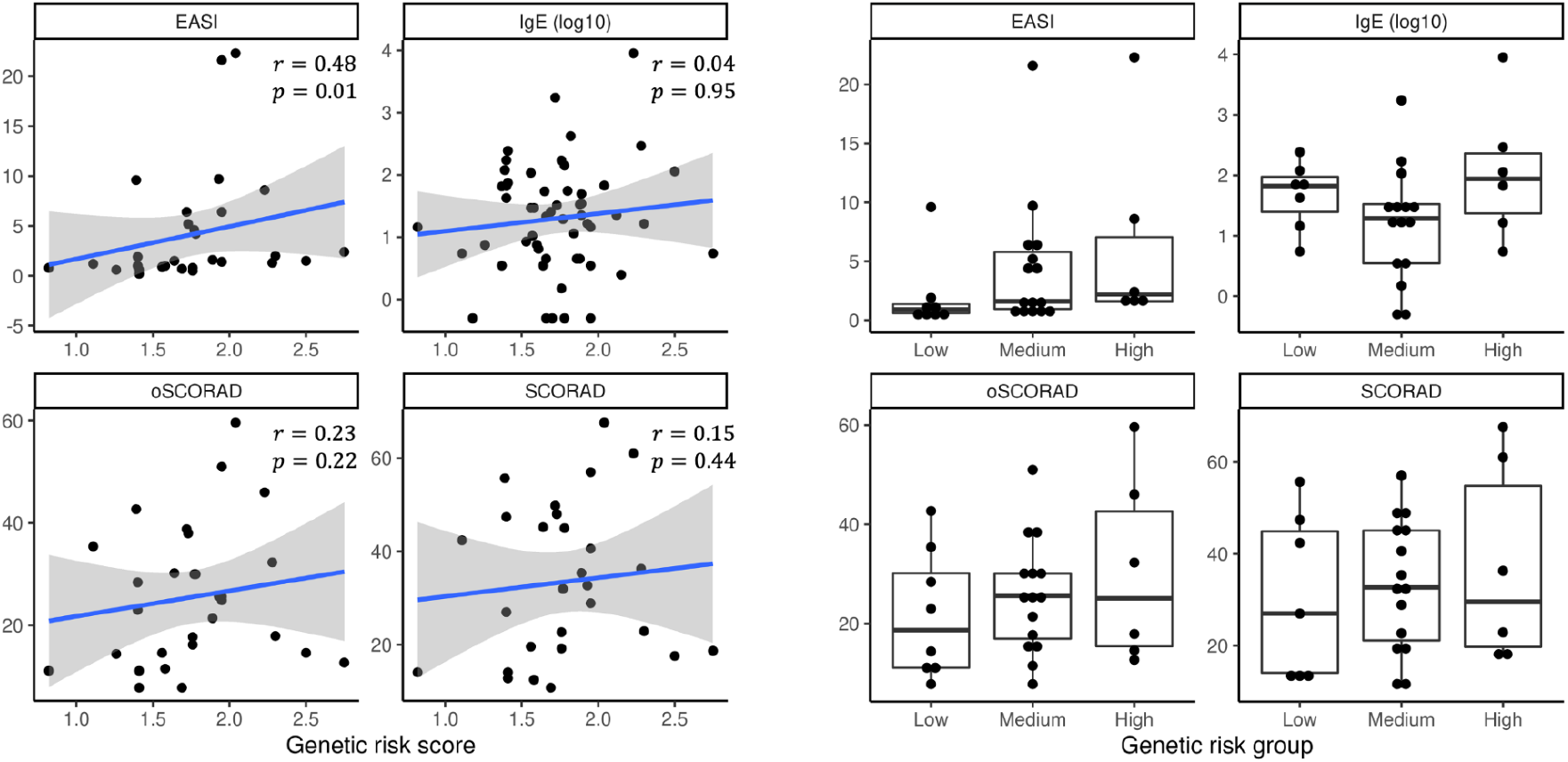
Genetic risk and clinical measurements. a) scatter plot showing the correlation between genetic risk score and clinical measurement, as well as the Spearman correlation coefficients and *p* values. b) Clinical measurements in different genetic risk groups.

**Figure S3.**
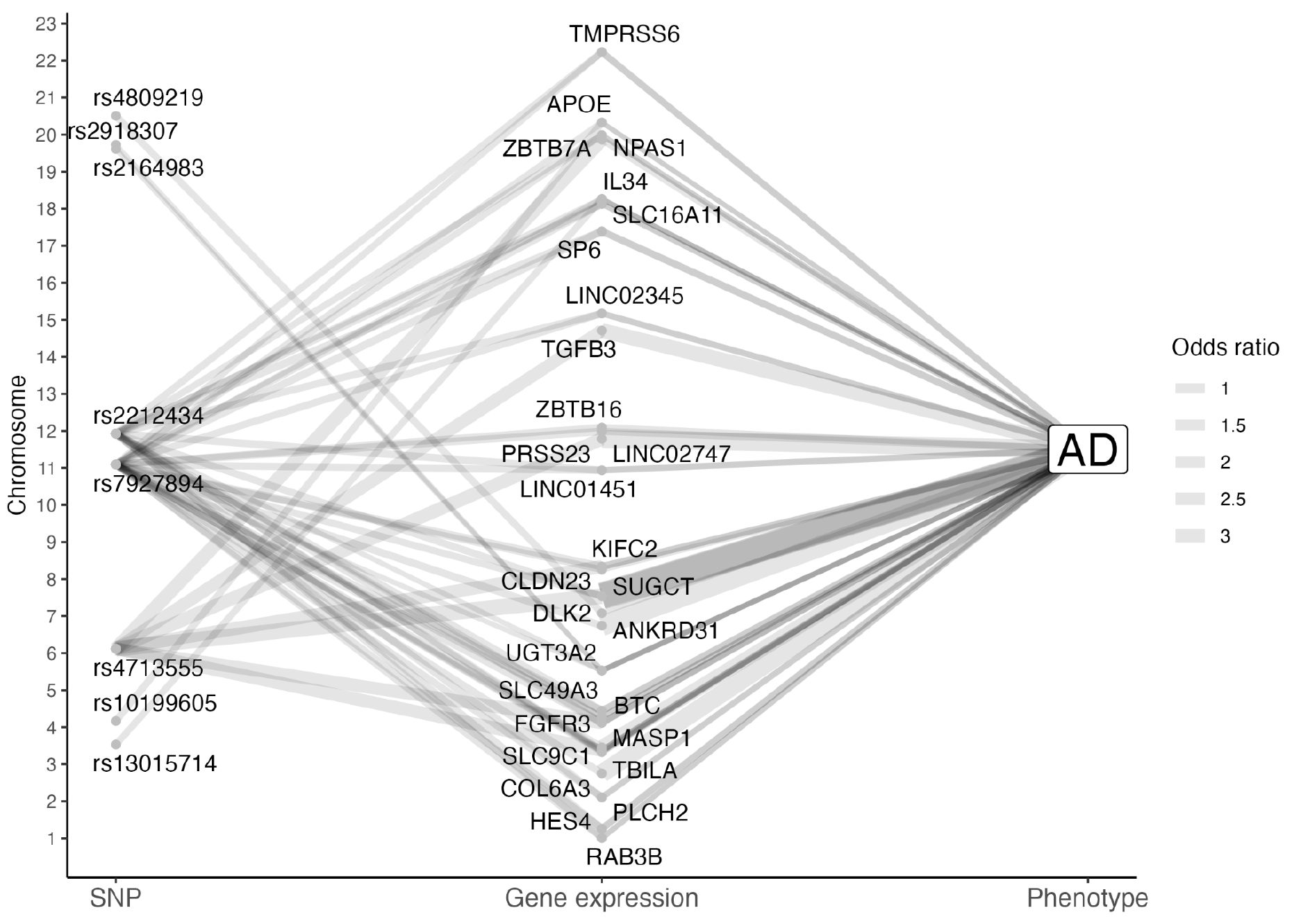
The interaction between SNPs, gene expression and AD as determined by two-parameter logistic regression models. The lines connecting SNP-gene expression, or gene expression-AD show significant interactions. The line width is proportional to the odds ratio.

